# UV1 cancer vaccine in pembrolizumab-treated patients with recurrent or metastatic PD-L1 positive head and neck squamous cell carcinoma: results from the randomized phase 2 FOCUS trial

**DOI:** 10.1101/2024.10.24.24315748

**Authors:** Anna Brandt, Konrad Klinghammer, Christoph Schultheiss, Lisa Paschold, Claudia Wickenhauser, Marcus Bauer, Anna Bergqvist, Dennis Hahn, Philippe Schafhausen, Mareike Tometten, Markus Blaurock, Henrike Barbara Zech, Chia-Jung Busch, Andreas Dietz, Urs Müller-Richter, Jürgen Alt, Andreas Boehm, Simone Kowoll, Jörg Steighardt, Alexander Lasch, Ingunn Hagen Westgaard, Marita Westhrin, Alexander Stein, Axel Hinke, Mascha Binder

## Abstract

**Background:** Human telomerase reverse transcriptase (hTERT) is highly expressed (75-100%) in head and neck squamous cell carcinoma (HNSCC). The FOCUS study examines the role of the hTERT-directed vaccine UV1 in combination with pembrolizumab in patients with recurrent or metastatic (R/M) HNSCC.

**Methods:** The FOCUS trial, a two-armed, open-label, non-comparative, randomized, multicenter phase 2 study, was designed to assess the efficacy and feasibility of UV1 as an add-on to pembrolizumab in the first-line treatment of patients with R/M PD-L1 positive HNSCC. A progression-free survival rate at 6 months (PFSR@6) of 40% was deemed promising for further development in a phase 3 setting. The trial was conducted in 10 centers in Germany.

**Results:** From August 2021 to July 2023, 25 patients were enrolled in the calibration arm A and 50 patients in the UV1 arm B. Median age was 65 years and 18% of patients had an ECOG performance score of 2. The PFSR@6 was 30% in the UV1 arm. No specific safety signals were observed in the UV1 arm apart from a reversible allergic reaction that appeared in one patient. At a median follow-up of almost one year (11.3 months), median overall survival was 13.1 months in the calibration arm A and 12.6 months in the UV1 arm B. Clinical trial identification number NCT05075122.

**Conclusions:** The addition of UV1 to pembrolizumab was safe but did not show an efficacy signal in this study population.

## Introduction

Head and neck squamous cell carcinoma (HNSCC) ranks as the seventh most prevalent cancer globally and is linked to tobacco use, alcohol consumption, and human papillomavirus (HPV) infection in oropharyngeal cancer (*1, 2*). Initial-stage therapy aims for a cure, yet more than half of HNSCC patients experience recurrent and/or metastatic (R/M) disease despite aggressive multimodal approaches (*3*). Many patients with R/M HNSCC qualify solely for palliative systemic therapy (*4*).

The incorporation of the epidermal growth factor receptor (EGFR) antibody cetuximab into platinum and fluorouracil (EXTREME regimen) substantially increased overall survival from 7.4 to 10.1 months as first-line treatment for R/M HNSCC, becoming the standard in 2008 (*4, 5*). More recently, the FDA approved pembrolizumab, a programmed death 1 (PD-1) immune checkpoint inhibitor, for first-line treatment based on the KEYNOTE-048 trial. Pembrolizumab with chemotherapy demonstrated improved overall survival compared to cetuximab with chemotherapy (median 13.0 vs. 10.7 months) (*6*). Particularly in patients with PD-L1 Combined Positive Score (CPS) ≥1 and CPS ≥20, pembrolizumab alone outperformed cetuximab with chemotherapy, underscoring increased efficacy with higher PD-L1 expression (*7*).

Despite durable responses in some patients, the majority (85-95%) of R/M HNSCC patients exhibit either no response or short-term benefit to immune checkpoint inhibitors (*3*). Insufficient T cell effector response might contribute to this lack of response (*8*). Efforts to enhance T cell response involve investigating therapeutic cancer vaccines, such as UV1, targeting human telomerase reverse transcriptase (hTERT), a key player in carcinogenesis activated in 85-90% of cancers (*8-12*). UV1 induced persistent immune responses lasting up to 7.5 years in phase I trials in patients with advanced melanoma, non-small cell lung cancer and prostate cancer, with promising results when combined with checkpoint inhibitors in the melanoma patients (*8*).

In HNSCC, where high hTERT expression (75-100%) is prevalent (*13*), the FOCUS trial (*14*) sought to evaluate the effectiveness of UV1 vaccination combined with pembrolizumab versus pembrolizumab alone in patients with PD-L1 positive R/M HNSCC.

## Results

### Disposition of study patients

Recruitment started in August 2021 and was closed in July 2023. Data base lock was July 24th, 2024. A total of 78 patients from a total of ten different study sites were randomized (Table 1). At its final closure, the database included information on 75 randomized, evaluable patients, 25 in arm A (calibration arm, pembrolizumab alone) and 50 in arm B (experimental arm, pembrolizumab and UV1 vaccination), forming the ITT full analysis set (Table 1). One patient did not receive the first protocol-defined pembrolizumab cycle and was excluded from the per-protocol (PP) population.

**Table 1:** Patient disposition and categories of evaluability (CONSORT)

| Parameter | Arm A | Arm B | Total |
| --- | --- | --- | --- |
| Randomised patients | 26 | 52 | 78 |
| Non-eligible for analysis (major primary protocol violation), randomised | - | 2* | 2* |
| No data available, randomised | 1** | - | 1** |
| Full analysis population (ITT) for this report*** | 25 | 50 | 75 |
| Evaluability (ITT) with respect to |  |  |  |
| Tumor response (iRECIST) at end of protocol treatment | 22 | 45 | 67*** |
| Progression-free survival (PFS) | 25 | 50 | 75 |
| **PFS rate at 6 months (primary endpoint)** | 25 | 50 | 75 |
| Overall survival (OS) | 25 | 50 | 75 |
| Evaluability for safety | 25 | 50 | 75 |

\* Two patients were excluded due to severe violation of entry criteria, present at randomization: performance status too poor (ECOG >2) and required antibiotic treatment; no protocol therapy applied.
\*\* Informed consent withdrawn one day after randomization; no protocol therapy applied. \*\*\* With a valid iRECIST re-staging.

### Baseline characteristics

Age and gender were equally distributed among the study arms, with a slight tendency to more elderly and male patients in the experimental arm (Table 2). In contrast, there was a slight trend towards less favourable ECOG performance status in arm A (Table 2).

**Table 2:** Baseline characteristics of eligible patients.

| Parameter | Arm A | Arm B | Total |
| --- | --- | --- | --- |
| n | 25 | 50 | 75 |
| **Age** |  |  |  |
| Mean ± SD | 64.5 ± 11.3 | 67.5 ± 10.5 | 66.5 ± 10.8 |
| **Sex** |  |  |  |
| Female | 7 (28%) | 8 (16%) | 15 (20%) |
| Male | 18 (72%) | 42 (84%) | 60 (80%) |
| **ECOG** |  |  |  |
| n | 24 | 48 | 72* |
| ECOG 0 | 8 (33%) | 17 (35%) | 25 (35%) |
| ECOG 1 | 9 (38%) | 24 (50%) | 33 (46%) |
| ECOG 2 | 7 (29%) | 6 (12%) | 13 (18%) |
| ECOG 3 | 0 (0%) | 1 (2%)** | 1 (1%)** |

\* The baseline vital sign visit was missing in 3 patients. \*\* Patient needing a wheelchair due to a non-oncological condition; accepted for inclusion by principal investigator; ECOG, Eastern Cooperative Oncology Group; n, number

### Disease status at recruitment

Overall, clinical features were relatively well balanced between the arms. Most patients (fewer in arm B) entered the FOCUS trial due to relapsing disease (Table 3). While advanced stages of the primary tumor were slightly more frequent in the UV1 arm B, more patients with distant metastases were randomised to arm A (Table 3). Distant metastases were predominantly located in the lung (Table 3). Median PD-L1 CPS was 35 (range 1-100) in arm A and 27 (range 1-100) in the UV1 arm B.

**Table 3:** Disease status at recruitment.

| Parameter | Arm A | Arm B | Total |
| --- | --- | --- | --- |
| **Disease status** |  |  |  |
| n | 25 | 50 | 75 |
| Relapse | 21 (84%) | 34 (68%) | 55 (73%) |
| Initial diagnosis | 4 (16%) | 16 (32%) | 20 (27%) |
| **Stage** |  |  |  |
| n | 25 | 47 | 72 |
| Stage I | - | 1 (2%) | 1 (1%) |
| Stage II | - | 5 (11%) | 5 (7%) |
| Stage III | 1 (4%) | 4 (9%) | 5 (7%) |
| Stage IVa | 6 (24%) | 12 (26%) | 18 (25%) |
| Stage IVb | 6 (24%) | 9 (19%) | 15 (21%) |
| Stage IVc | 12 (48%) | 16 (34%) | 28 (39%) |
| **Location of metastases** |  |  |  |
| n | 15 | 22 | 37 |
| Bone | 1 (7%) | 2 (9%) | 3 (8%) |
| Liver | 4 (27%) | 3 (14%) | 7 (19%) |
| Lung | 8 (53%) | 12 (55%) | 20 (54%) |
| Skin | 2 (13%) | 1 (5%) | 3 (8%) |
| Other | 4 (27%) | 6 (27%) | 10 (27%) |
| **Pre-treatment** |  |  |  |
| n | 25 | 50 | 75 |
| Surgery | 13 (52%) | 29 (58%) | 42 (56%) |
| Adjuvant radiotherapy | 3 (12%) | 7 (14%) | 10 (13%) |
| Adjuvant chemoradiotherapy | 8 (32%) | 20 (40%) | 28 (37%) |
| Definitive chemoradiotherapy | 3 (12%) | 8 (16%) | 11 (15%) |
| Radiotherapy | 5 (20%) | 7 (14%) | 12 (16%) |
| Other | 2 (8%) | 3 (6%) | 5 (7%) |

### Antineoplastic treatment and duration of follow-up

Median duration of pembrolizumab treatment was 15.1 weeks (range 0.1-98.6) in arm A versus 12.1 weeks (range 0-84.3) in arm B. Median number of UV1 administrations per patient was 8 (range 1-8). About half of the patients (54%) received the maximum number of vaccine administrations.

Mean duration of follow-up since randomization was slightly shorter than one year (11.3 months), ranging up to 28.7 months, and was quite similar in both study arms (11.7 months in arm A and 11.1 months in arm B).

### Tumor response

Tumor response according to iRECIST as investigated at the end of protocol therapy, is shown in Table 4. A total of 67 out of 75 patients had a valid restaging result at this time point. The corresponding overall response rate (CR + PR = ORR) was 45% in arm A (95% CI, 24%-68%) and 22% in arm B (95% CI, 11%-37%) (p = 0.086).

**Table 4:** Response at the end of protocol therapy.

| Category | Arm A | Arm B | Total |
| --- | --- | --- | --- |
| n | 22 | 45 | 67 |
| CR | 3 (14%) | 2 (4%) | 5 (7%) |
| PR | 7 (32%) | 8 (18%) | 15 (22%) |
| SD | 1 (5%) | 6 (13%) | 7 (10%) |
| PD unconfirmed | 4 (18%) | 12 (27%) | 16 (24%) |
| PD confirmed | 7 (32%) | 17 (38%) | 24 (36%) |

### Progression-free survival

The results for the primary endpoint of the study, i.e. the crude proportion of patients surviving without progression at 6 months after randomization, are shown in Table 5. With respect to the formal study hypothesis, the two-sided 80% CI (corresponding to the 90% one-sided CI as relevant for superiority) of the rate in the experimental arm B does not exclude the 25% boundary, which was pre-defined as futility threshold. Moreover, the 40% finding in the group treated with pembrolizumab alone (arm A) is higher than expected. Thus, a positive signal for the experimental treatment cannot be derived from the results of this phase 2 study.

**Table 5:**
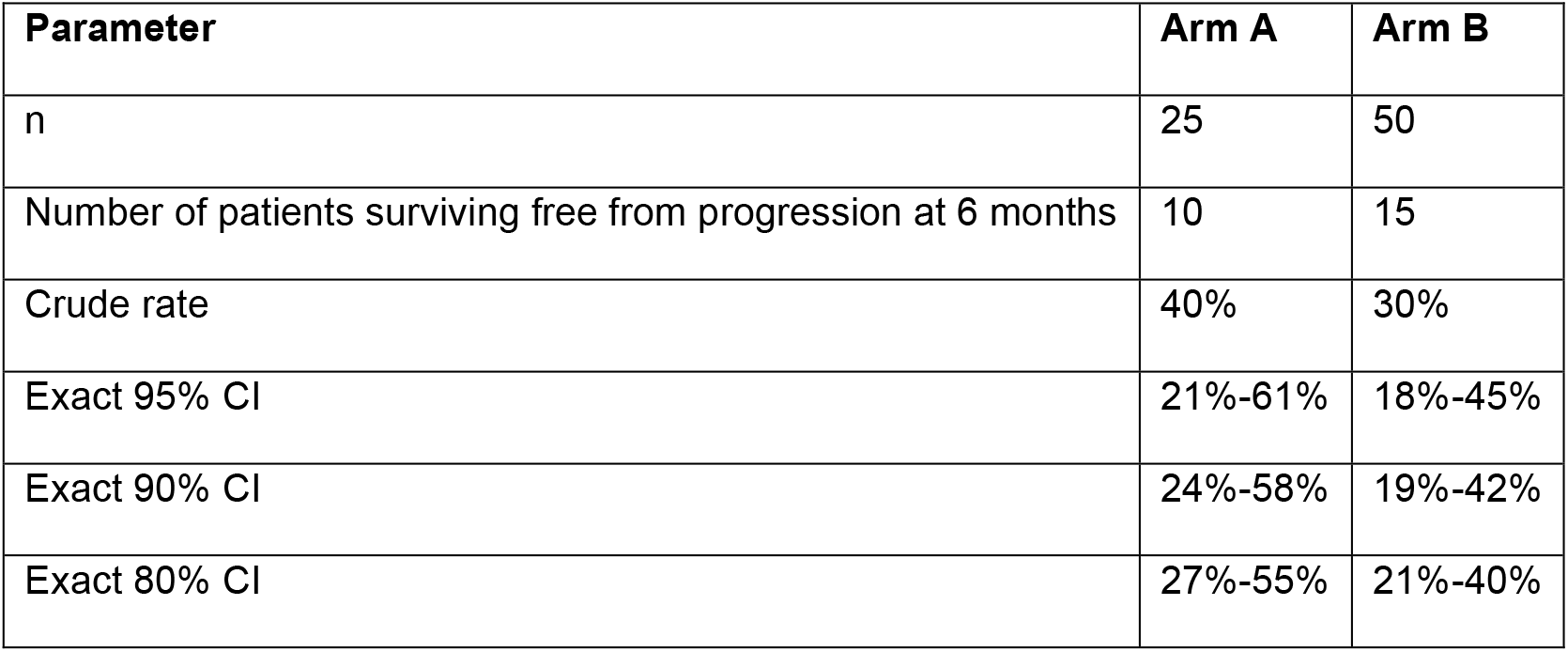
Crude PFS rate at 6 months (primary endpoint)

Correspondingly, the Kaplan-Meier estimation of progression-free survival (PFS) from the time point of randomization (Figure 1A), based on a total of 63 observed PFS events in the ITT population of 75 patients (84%), shows no major difference between the arms. The medians in arm A and B are similar with 3.9 (95% CI, 3.0-13.1) and 3.3 months (95% CI, 3.2-4.4), respectively. The 6-month PFS rates estimated according to Kaplan-Meier are 42% (95% CI, 26%-67%) and 31% (95% CI, 20%-47%), respectively. The hazard ratio (HR) amounts to 1.3 (95% CI, 0.76-2.23) suggesting no relevant difference between the arms.

**Figure 1.**
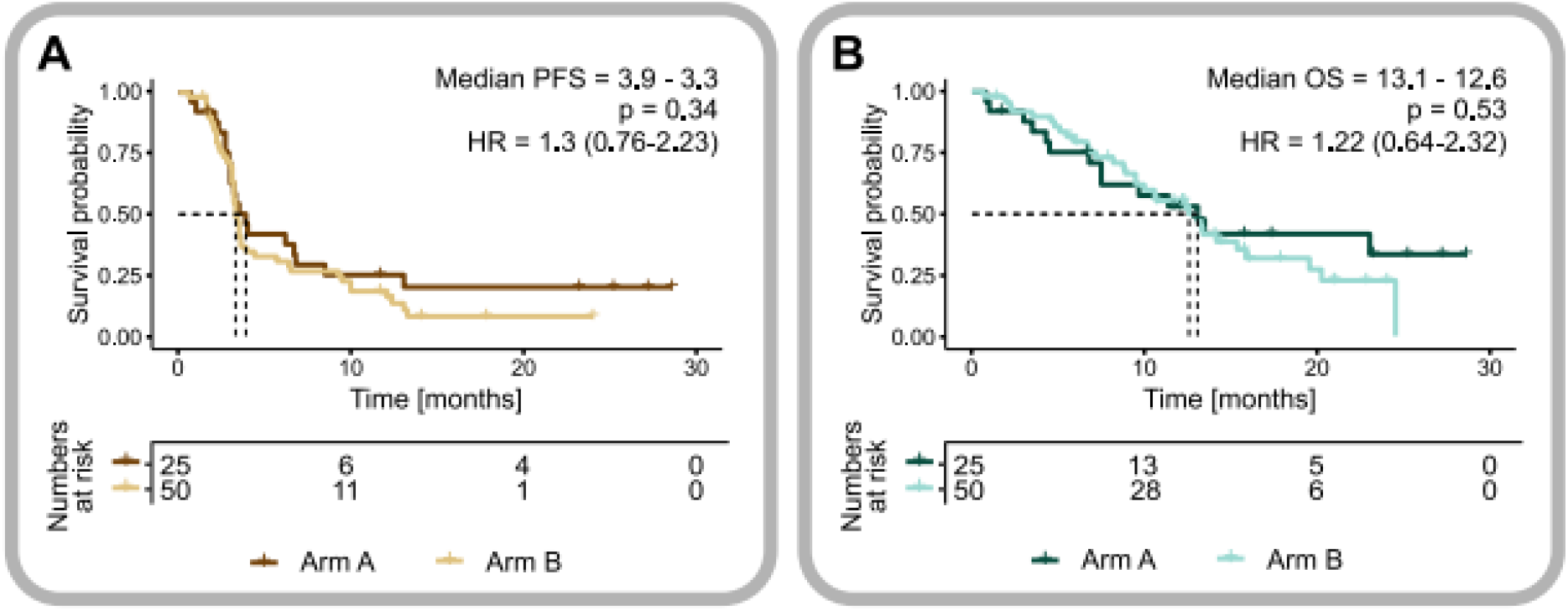
Efficacy of UV1 + pembrolizumab treatment in patients with R/M HNSCC. **A**, Kaplan-Meier estimates of progression-free survival. **B**, Kaplan-Meier estimates of overall survival.

The Kaplan-Meier estimation of overall survival (OS) from the time point of randomization, based on a total of 46 observed deaths in the ITT population of 75 patients (61%) is provided in Figure 1B. The medians in arm A and B were similar with 13.1 (95% CI, 7.7-undefined) and 12.6 months (95% CI, 9.5-19.6), respectively. The hazard ratio amounts to 1.22 (95% CI: 0.64-2.32).

### Safety

The safety analysis is based on the total population of 75 patients. A total of 61 grade 3 or higher adverse events (AEs) were recorded in the ITT population, 44% of patients in arm A and 50% in arm B experienced at least one such AE. Table 6 presents an overview of observed adverse events, overall and according to severity and other pre-defined categories. No relevant differences between the two study arms were detected.

**Table 6:** Overview of adverse events.

| Parameter | Arm A | Arm B |
| --- | --- | --- |
| n | 25 | 50 |
| Total number of adverse events reported | 124 | 263 |
| Number of adverse events of severity grade 3 to 5 | 21 | 40 |
| Number of patients with any adverse event | 25 (100%) | 47 (94%) |
| Number of patients with any adverse event of severity grade 3 to 5 | 11 (44%) | 25 (50%) |
| Number of patients with any serious AE (SAE) | 10 (40%) | 28 (56%) |
| Number of patients with any AE of special interest | NA | 1 (2%)* |

\*Severe anaphylactic reaction

Grade 3 or higher AEs are summarized in Table 7.

**Table 7.** Patients experiencing grade 3 or higher adverse events.

| Adverse event | Arm A<br>n=25 | Arm B<br>n=50 |
| --- | --- | --- |
| Infection | 5 (20%) | 7 (14%) |
| Sepsis | 1 (4%) | 0 |
| Lipase increased | 0 | 1 (2%) |
| Abdominal pain | 0 | 1 (2%) |
| Diarrhea | 0 | 1 (2%)* |
| Allergic reaction | 0 | 1 (2%) |

\*Grade missing

### hTERT tissue expression

hTERT tissue expression at baseline was assessed in 63 patients. There was a trend to longer progression free survival with hTERT expression >0 in arm B, however, this trend was also seen in the control arm A (Figure 2).

**Figure 2.**
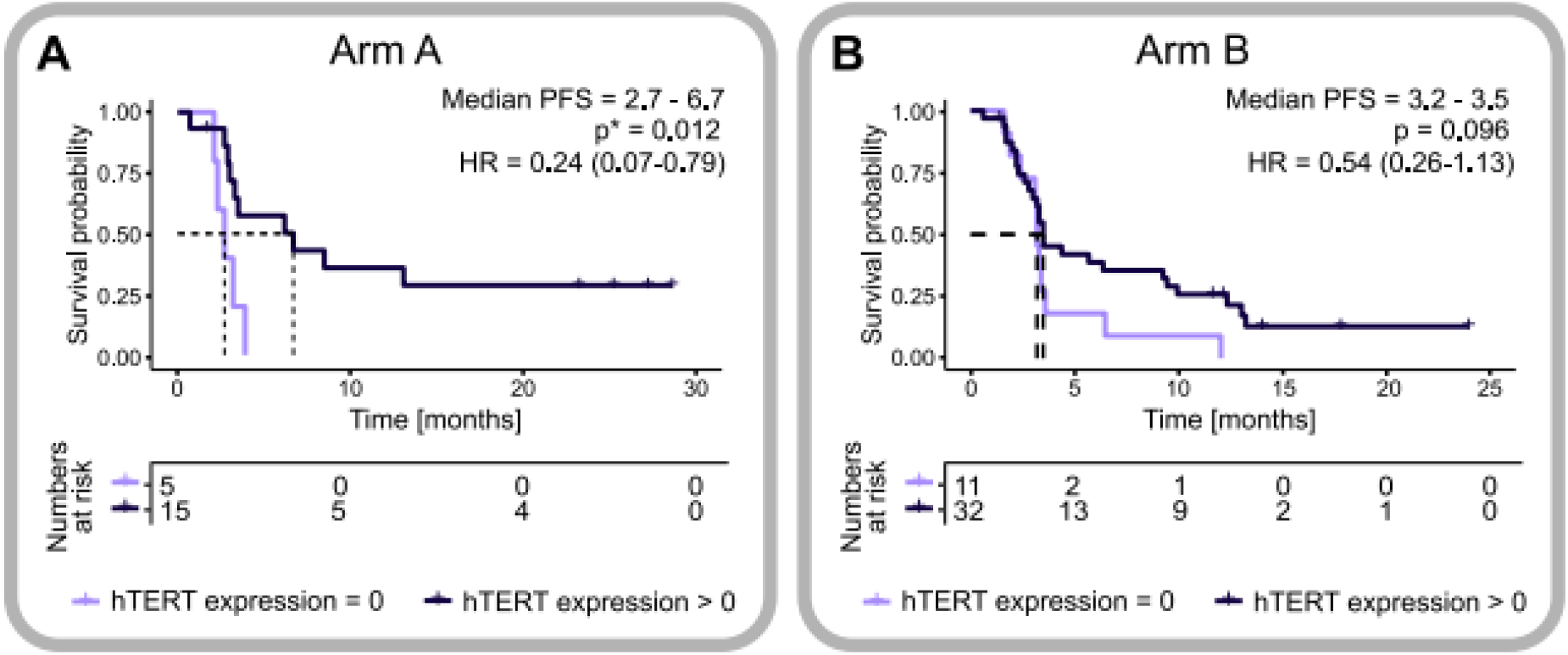
Kaplan-Meier estimates of progression-free survival of patients with hTERT expression >0 and 0 in arm A (**A**), and arm B (**B**).

### Immune response to UV1

To assess UV1-specific T cell responses over time, IFN-γ secretion was quantified via ELISPOT assays after stimulation of patient-derived peripheral blood mononuclear cells (PBMCs) obtained at visit 1 (baseline, BL) and follow-up (FU) timepoints (visit 6 and/or end of treatment (EOT)/progressive disease (PD)) with UV1 vaccine peptides (p719-20, p728, p725 or the UV1 peptide pool). As shown in Figure 3, individual patients in both study arms displayed UV1-directed T cell responses at baseline. Notably, UV1-directed T cell responses were almost undetectable in the FU time points in arm A without vaccination (Figure 3). UV1-directed T cell responses were observed in 12 of 14 vaccinated individuals (Figure 3, arm B). The highest reactivity was detected after stimulation with the UV1 peptide pool or the p719-20 peptide, while reactivity to the single hTERT peptides p728 and p725 was substantially lower (Figure 3).

**Figure 3:**
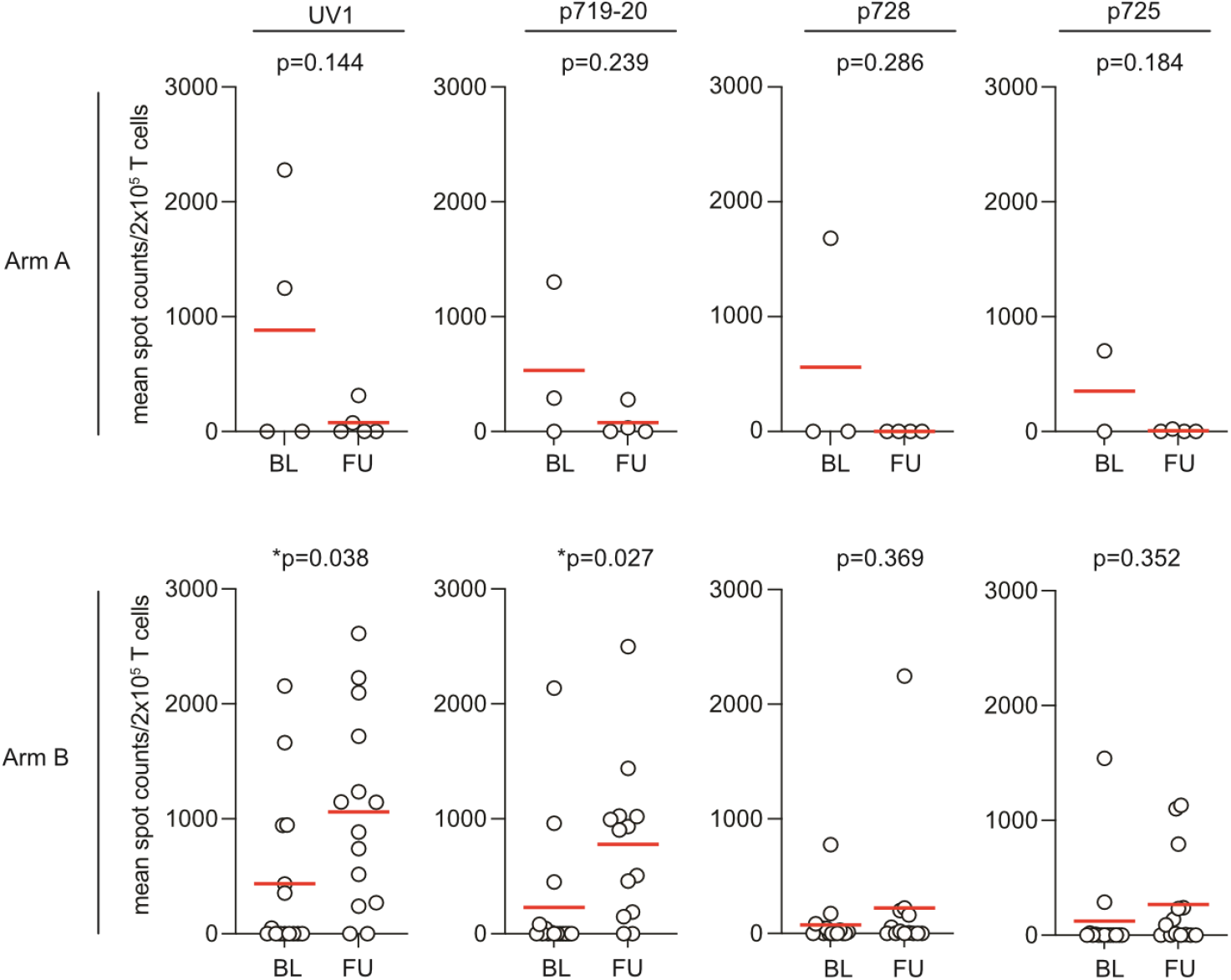
Quantification of pre- and post-vaccination immune responses to UV1. UV1-directed T cell responses were quantified using IFN-γ ELISPOT after peptide stimulation of patient derived PBMCs. Mean ELISPOT counts per patient are shown for both arms. Sample numbers in arm A: BL n=4, FU n=5; arm B: BL n=16, FU n=13. BL sampled at visit 1, FU sampled at visit 6 and/or end of treatment (EOT)/progressive disease (PD). UV1 refers to hTERT peptide mix (=UV1 vaccine) encompassing the peptides p719-20 (hTERT amino acids 660-689, peptide 728 (hTERT amino acids 651-665) and peptide 725 (hTERT amino acids 691-705).

## Discussion

The treatment of advanced or recurrent head and neck cancer remains a significant challenge. Current treatment options, such as chemotherapy and immunotherapy are established treatment options albeit with limited efficacy (*5, 6, 15*). Moreover, most pivotal trials excluded more frail patients who are likely to experience severe side effects and have poorer overall outcomes (*16*). Therefore, trial results are often not representative for patients with head and neck cancer many of which suffering from concomitant medical conditions (*17*). Safe and at the same time effective treatment options that spare toxicities in these frail patients would represent a significant advancement.

To address these unmet needs, we conducted a phase 2 trial evaluating the hTERT vaccine UV1 in combination with pembrolizumab in a cohort of patients with advanced head and neck cancer. We enrolled patients up to an ECOG performance status of 2. Unfortunately, our findings indicate that the hTERT vaccine UV1 did not demonstrate a signal of efficacy in this population.

Our results stand in contrast to those of previous studies using the same vaccine in other cancers, such as melanoma, where more consistent immune responses were observed (*8*). In the recently published NIPU-trial a positive overall survival signal was observed in mesothelioma patients when UV1 was combined with ipilimumab and nivolumab (*18*). This discrepancy suggests that the immune environment and tumor biology of head and neck cancers may differ significantly from other tumor types, thereby influencing the efficacy of immunotherapeutic approaches such as UV1.

Despite these disappointing results, our study provides valuable insights into the efficacy of pembrolizumab in a more real-world patient population. In contrast to the KEYNOTE-048 trial, which limited the inclusion criteria to an ECOG of 1 due to the possible administration of additional chemotherapy, the FOCUS trial enrolled patients with an ECOG up to 2, made possible by exclusive use of pembrolizumab (*6*). In this frailer patient population overall response rate, progression-free survival and overall survival with pembrolizumab were comparable to the outcomes of PD-L1 positive patients in the KEYNOTE-048 trial (*19*). Recently, the results of the randomized phase 3 trial ELAN UNFIT have been published, which compared the efficacy and safety of cetuximab to methotrexate in older, frail patients with R/M HNSCC (*16*). Even though this trial did not show an improvement in failure-free survival with cetuximab versus methotrexate, more trials such as ELAN UNFIT or FOCUS are needed to better understand the unique therapeutic challenges and needs of older patients with head and neck cancer (*20*).

Moreover, our study contributes to the debate about the limitations of current immunotherapy approaches in advanced head and neck cancers. The lack of efficacy observed with the hTERT vaccine UV1 raises important questions about the role of telomerase-targeted immunotherapy in this setting and emphasizes the need for continued exploration of alternative strategies. Our results suggest that while telomerase-targeted immunotherapy holds promise for certain cancers, its application in head and neck cancers does not seem to be promising or would need other therapeutic modalities to enhance efficacy.

## Methods

### Clinical trial

Main inclusion criteria consisted of: Patients had to be at least 18 years of age, ECOG-performance score 0-2, with histologically confirmed diagnosis of non-resectable recurrent or metastatic head and neck squamous cell carcinoma with at least one measurable tumor lesion as per RECIST v.1.1. Patients had to be eligible for pembrolizumab monotherapy with a PD-L1 CPS ≥1 and adequate laboratory parameters. The trial was conducted at 10 centers in Germany in compliance with the Declaration of Helsinki. The study protocol was approved by the local ethics committees and authorized by the competent authority. All participants provided written informed consent. The trial is registered with ClinicalTrials.gov (NCT05075122).

Eligible patients were randomized to pembrolizumab mono (arm A) or pembrolizumab in combination with UV1 vaccination (arm B). Randomization was done via an online tool (secuTrial®) of the Coordination Center for Clinical Trials, Medical Faculty, Martin-Luther-University Halle-Wittenberg, Halle (Saale), Germany. All patients received pembrolizumab until disease progression or up to two years. In Arm A patients received pembrolizumab 200 mg i.v. every 3 weeks, administration of pembrolizumab started in week 1 (treatment duration of 12 weeks). In arm B patients received pembrolizumab 200 mg i.v. every 3 weeks in combination with UV1 vaccination (300 µg UV1 s.c. in addition to 75 µg GM-CSF s.c.). Three UV1 doses were applied during week 1, followed by 5 vaccinations every 3 weeks on day 1 of each cycle. In arm B administration of pembrolizumab started at week 2, one week later than in arm A (treatment duration of 13 weeks).

Radiological baseline assessment by computed tomography of the neck, chest, abdomen and pelvis not older than 4 weeks before randomization was performed. Patients were assessed on week 1, day 1 (visit 1), week 1, day 3 (visit 2), week 1, day 5 (visit 3), week 2 (visit 4), week 5 (visit 5) week 8 (visit 6), week 11 (visit 7), week 14 (visit 8), at the end of treatment (EOT), and at progressive disease (PD) if applicable. All patients were evaluated every 3 months after EOT until death or maximal 12 months after last patient first visit.

The legal funder (sponsor) of the trial was the University Medical Center Halle (Saale), Germany.

### Statistical analysis

The primary endpoint was PFSR@6, defined as the proportion of patients alive without progression at six months after randomization divided by the total number of evaluable patients, and the low boundary of its one-sided 90% confidence interval, corresponding to a type I error level of 0.1. To detect a promising efficacy level of PFSR@6 ≥40% against a futility level of ≤25%, as expected for pembrolizumab single-agent treatment, with a power of 80%, 46 evaluable patients were required in the experimental arm (*21*). By 2:1 ratio, patients were allocated to a randomized reference arm, to allow some control of selection bias.

Due to the lack of adequate power, statistical comparisons between the study arms must be considered exploratory, with all p values being two-sided. Fisher’s exact test was applied for nominal data, Student’s t test for continuously distributed data, and the log rank test for time-to-event analysis, with hazard ratios derived from Cox models.

### Immunohistochemistry (IHC) for TERT

IHC staining was performed on a Bond III automated immunostainer (Leica Biosystems Nussloch GmbH, Wetzlar, Germany) using the Bond Polymer Refine Detection Kit (DS9800-CN). The polyclonal anti-TERT antibody (Telomerase catalytic subunit Antibody, 600-401-252, Rockland Immunochemicals Inc., Limerick, PA, USA) was applied in a dilution of 1:500. The staining intensity was scored using the H-score as described elsewhere (*22*).

### Quantification of anti-hTERT immune responses

PBMCs were seeded in 48-well plates at 6 million cells per well in IMDM medium (Gibco) supplemented with 10% heat-inactivated human serum, penicillin/streptomycin and 50 µM beta-mercaptoethanol. The PBMCs were stimulated with UV1 drug product (peptide 725; hTERT 691-705 (RTFVLRVRAQDPPPE), peptide 719-20; hTERT 660-689 (ALFSVLNYERARRPGLLGASVLGLDDIHRA), peptide 728; hTERT 651-665 (AERLTSRVKALFSVL) (Corden Pharma, Caponago, Italy) at a concentration of 15 μM each and 20 µg/ml poly-ICLC (Oncovir). Medium containing 20 U/ml IL-2 was replaced every two days. For IFN-γ ELISPOT assays, cells were harvested on day 12, seeded as triplicates at 0.1 million each in ELISPOT plates (Human IFN-γ ImmunoSpot, CTL). The UV1 peptides were added at 20 μM each or 20 µM UV1 peptide mix (= UV1 cancer vaccine). T cells alone served as negative control, T cells stimulated with 0.08μg/ml SEC-3 superantigen (Toxin Technology Inc. Sarasota, FL, USA) as positive control. Cells were incubated for 22 hours before enumeration of the detected spots using an automated analyzer, CTL IMMUNOSPOT S5 VERSA-02-9030 (Cellular Technology Ltd, Shaker Heights, OH, USA). Specific spots were calculated by subtracting the mean number of spots detected in the medium-only control from the mean number of spots detected in the experimental samples.

## Contributors

Concept and study design: MBi, AS, CJB, MW, JS, IHW, SK, AL, HBZ; Patient recruitment, data collection: KK, MBi, DH, PS, MT, MBl, JA, AD, UMR, ABo; Translational study design, molecular studies, data collection, data analysis, data interpretation: ABr, CS, LP, KK, CW, MBa, AH, MBi, MW, ABe, IHW; Statistical analysis: AH, LP; Drafting of the manuscript: ABr, MBi, CS. Critical revision of the manuscript for important intellectual content: all authors. All authors agreed to submit the manuscript and read and approved its content.

## Data Availability

All data produced in the present study are available upon reasonable request to the authors.

## Acknowledgements

We thank all patients and families as well as all participating study centers.

## Notes

**Conflicts of interest:** Mascha Binder received institutional research grants from Merck, BMS, Hexal, Novartis, German Cancer Aid (Krebshilfe), German Research Foundation and the Federal Ministry of Education and Research as well as honoraria for lectures and advisory board meetings by Celgene, Janssen, Gilead, Merck, Roche, Amgen, Sanofi-Aventis and BMS. She received funding for the FOCUS trial from ULTIMOVACS. Alexander Stein received research funding from MSD and serves as an advisory board member for MSD. Urs Müller-Richter serves as a consultant or advisor and/or received honoraria from AstraZeneca, BioNTech, BMS, KuraOncology, Merck, MSD, Novartis, and Sanofi. Jürgen Alt serves as an advisor for AstraZeneca, MSD, Novartis, Roche, BMS, Janssen, and Merck and received honoraria from AstraZeneca, BMS, Roche, and Boehringer Ingelheim. Konrad Klinghammer serves as a consultant or advisor and/or received honoraria from MSD, Merck, BMS, Roche, Novartis, Sanofi, Bayer, BioNTech, Boehringer Ingelheim, and onkowissen. Anna Brandt received honoraria from Merck. Dennis Hahn serves as a consultant or advisor and/or received honoraria from BMS, MSD and Merck. Henrike Barbara Zech serves as a consultant and /or received honoraria from MSD, Merck, BMS, Sanofi, Regeneron. Chia-Jung Busch received institutional research grants from BMS and German Cancer Aid (Krebshilfe) as well as honoraria for lectures and advisory board meetings by Bayer, BMS, GSK, Merck, MSD, Sanofi-Aventis. Ingunn Hagen Westgaard, Anna Bergqvist and Marita Westhrin are employed by Ultimovacs ASA. Philippe Schafhausen serves as a consultant or advisor and/or received honoraria from Alexion, AOP Orphan Pharmaceutical, Blueprint Medicines, GSK, MSD, Merck, BMS, Novartis, Pfizer, Roche, Sanofi, and Sobi. All other authors declare no potential conflict of interest.

### Competing Interest Statement

Mascha Binder received institutional research grants from Merck, BMS, Hexal, Novartis, German Cancer Aid (Krebshilfe), German Research Foundation and the Federal Ministry of Education and Research as well as honoraria for lectures and advisory board meetings by Celgene, Janssen, Gilead, Merck, Roche, Amgen, Sanofi-Aventis and BMS. She received funding for the FOCUS trial from ULTIMOVACS. Alexander Stein received research funding from MSD and serves as an advisory board member for MSD. Urs Mueller-Richter serves as a consultant or advisor and/or received honoraria from AstraZeneca, BioNTech, BMS, KuraOncology, Merck, MSD, Novartis, and Sanofi. Juergen Alt serves as an advisor for AstraZeneca, MSD, Novartis, Roche, BMS, Janssen, and Merck and received honoraria from AstraZeneca, BMS, Roche, and Boehringer Ingelheim. Konrad Klinghammer serves as a consultant or advisor and/or received honoraria from MSD, Merck, BMS, Roche, Novartis, Sanofi, Bayer, BioNTech, Boehringer Ingelheim, and onkowissen. Anna Brandt received honoraria from Merck. Dennis Hahn serves as a consultant or advisor and/or received honoraria from BMS, MSD and Merck. Henrike Barbara Zech serves as a consultant and /or received honoraria from MSD, Merck, BMS, Sanofi, Regeneron. Chia-Jung Busch received institutional research grants from BMS and German Cancer Aid (Krebshilfe) as well as honoraria for lectures and advisory board meetings by Bayer, BMS, GSK, Merck, MSD, Sanofi-Aventis. Ingunn Hagen Westgaard, Anna Bergqvist and Marita Westhrin are employed by Ultimovacs ASA. Philippe Schafhausen serves as a consultant or advisor and/or received honoraria from Alexion, AOP Orphan Pharmaceutical, Blueprint Medicines, GSK, MSD, Merck, BMS, Novartis, Pfizer, Roche, Sanofi, and Sobi. All other authors declare no potential conflict of interest.

### Clinical Trial

Clinical trial identification number NCT05075122

### Funding Statement

The trial including translational research was funded by a research grant from ULTIMOVACS.

### Author Declarations

The trial was conducted at 10 centers in Germany in compliance with the Declaration of Helsinki. All participants provided written informed consent. Ethical approval was given by all of the following Ethics committees: Ethics Committee of the Medical Faculty of Martin Luther University Halle-Wittenberg, Halle Saale, Germany Ethics Committee at the Medical Faculty of Leipzig University, Leipzig, Germany Ethics Committee of the State Medical Association, Mainz, Germany Ethics Committee at the Medical Faculty of RWTH Aachen, Aachen, Germany Ethics Committee at the State Medical Association of Baden-Wuerttemberg, Stuttgart, Germany Ethics Committee of the State of Berlin, Berlin, Germany Ethics Committee of the Medical Association Hamburg, Hamburg, Germany Ethics Committee of the Medical Faculty of the University of Wuerzburg, Wuerzburg, Germany Ethics Committee at the University Medical Center Greifswald, Institute of Pharmacology, Greifswald, Germany Ethics Committee of the Saxon State Medical Association, Dresden, Germany

